# COVID-19 screening strategies that permit the safe re-opening of college campuses

**DOI:** 10.1101/2020.07.06.20147702

**Authors:** A. David Paltiel, Amy Zheng, Rochelle P. Walensky

## Abstract

**Importance:** The COVID-19 pandemic poses an existential threat to many US residential colleges: either they open their doors to students in September or they risk serious financial consequences.

**Objective:** To define SARS-CoV-2 screening performance standards that would permit the safe return of students to campus for the Fall 2020 semester.

**Design:** Decision and cost-effectiveness analysis linked to a compartmental epidemic model to evaluate campus screening using tests of varying frequency (daily-weekly), sensitivity (70%-99%), specificity (98%-99.7%), and cost ($10-$50/test). Reproductive numbers R_t_ = {1.5, 2.5, 3.5} defined three epidemic scenarios, with additional infections imported via exogenous shocks. We generally adhered to US government guidance for parameterization data.

**Participants:** A hypothetical cohort of 5000 college-age, uninfected students.

**Main Outcome(s) and Measure(s):** Cumulative tests, infections, and costs; daily isolation dormitory census; incremental cost-effectiveness; and budget impact. All measured over an 80-day, abbreviated semester.

**Results:** With R_t_ = 2.5, daily screening with a 70% sensitive, 98% specific test produces 85 cumulative student infections and isolation dormitory daily census averaging 108 (88% false positives). Screening every 2 (7) days nets 135 (3662) cumulative infections and daily isolation census 66 (252) with 73% (4%) false positives. Across all scenarios, test frequency exerts more influence on outcomes than test sensitivity. Cost-effectiveness analysis selects screening every {2, 1, 7} days with a 70% sensitive test as the preferred strategy for R_t_ = {2.5, 3.5, 1.5}, implying a screening cost of {$470, $920, $120} per student per semester.

**Conclusions & Relevance:** Rapid, inexpensive and frequently conducted screening – even if only 70% sensitive – would be cost-effective and produce a modest number of COVID-19 infections. While the optimal screening frequency hinges on the success of behavioral interventions to reduce the base severity of transmission (R_t_), this could permit the safe return of student to campus.

**KEY POINTS:** *Question:* What SARS-CoV-2 screening and isolation program will keep U.S. residential college students safe and permit the reopening of campuses?

*Findings:* Frequent screening (every 2 or 3 days) of all students with a low-sensitivity, high-specificity test will control outbreaks with manageable isolation dormitory utilization at a justifiable cost.

*Meaning:* Campuses can safely reopen in the Fall 2020 but success hinges on frequent screening and uncompromising, continuous attention to basic prevention and behavioral interventions to reduce the baseline severity of transmission.

## INTRODUCTION

Universities across the United States are struggling with the question of whether and how to reopen for the Fall 2020 semester.^1,2^ Residential colleges – with their communal living arrangements, shared dining spaces, intimate classrooms, and a population of young adults anxious to socialize – pose a particular challenge. In the absence of an effective vaccine, a proven therapy, and/or sufficient herd immunity, the best hope for re-opening campuses in the fall is likely to be a robust strategy of behavior-based prevention combined with regular monitoring to rapidly detect, isolate, and contain new SARS-CoV-2 infections, when they occur.^3^

Evidence on the available monitoring technologies and their performance is limited and rapidly evolving. The FDA is currently evaluating over 100 candidate tests for the presence of SARS-CoV-2 infection or antibodies.^4,5^ The uncertainties span a broad range, including the logistics of deployment, the ease and comfort of sample collection, and the accuracy, scalability, turn-around-time and cost of test kits. After a new COVID-19 case is detected, further questions emerge regarding how to conduct subsequent tracing, how to isolate detected cases in the context of congregate housing arrangements, and how to protect other at-risk populations, including faculty, staff, and members of the surrounding community.^6^ These uncertainties underscore the pressing need for both a generalized assessment of population-wide screening for SARS-CoV-2 and a comprehensive plan for university reopening.

For many U.S. colleges, COVID-19 poses an existential threat: either they open their doors to students in September or they suffer severe financial consequences.^7^ University administrators struggling with this dilemma must nevertheless keep in mind that their first priority is the safety of the students in their care. In this paper, we offer specific recommendations on the design of a virologic monitoring program that will keep students safe at an affordable cost. Our specific research objectives are: first, to define the minimum performance attributes of a SARS-CoV-2 monitoring program (e.g., its frequency, sensitivity, specificity, and cost) that could ensure that college students are kept safe; second, to understand how those minimum performance standards might change under varying assumptions about the severity of the epidemic and the success of behavioral and social distancing interventions; third, to suggest what isolation and treatment capacity would need to be in place; and finally, to forecast what all this might cost and to help decision makers make sense of that information to address the question of a screening and monitoring program’s “value.”

## METHODS

### Study Design

We adapted a simple compartmental epidemic model to capture the essential features of the situation facing university decision makers: the epidemiology of SARS-CoV-2; the natural history of COVID-19 illness; and regular mass screening to detect, isolate, and contain the presence of SARS-CoV-2 in a residential college setting (**Figure S1**). A spreadsheet implementation of the model permitted us to vary critical epidemic parameters and to examine how different test performance attributes (frequency, sensitivity, specificity, cost) would translate into outcomes. Model input data (**Table 1**) were obtained from a variety of published sources, adhering whenever possible to data guidance for modelers recently issued by the Centers for Disease Control and Prevention (CDC) and the Office of the Assistant Secretary for Preparedness and Response (ASPR).^8-18^ We defined three increasingly pessimistic epidemic scenarios and estimated both cumulative outcomes (e.g., tests administered; true/false positives; new infections; and person-days requiring isolation) and economic performance (e.g., costs, incremental cost-effectiveness, and budget impact) over an abbreviated 80-day semester, running from Labor Day through Thanksgiving.^2^ We assumed a medium-sized college setting with a target population of 5000 students, all of them <30 years old and non-immune, living in a congregate setting.^18,19^ We “seeded” this population with 10 undetected, asymptomatic cases of SARS-CoV-2 infection.

**Table 1.** Model input parameters and scenarios.

| Model inputs |  |  |  | References |
| --- | --- | --- | --- | --- |
| Compartments | Initial population size (n) |  |  |  |
| Non-infected, susceptible | 4,990 |  |  | 18 |
| Infected, asymptomatic | 10 |  |  | Assumption |
| All other compartments | 0 |  |  | Assumption |
| Time horizon (days) | 80 |  |  | 2 |
| Disease dynamics |  |  |  |  |
| Time to recovery (1/ρ) | 14 days |  |  | 9,10 |
| Time to false positive return (1/μ) | 1 day |  |  | Assumption |
| Probability of symptoms given infection (%) | 30 |  |  | 11-13 |
| Symptomatic case fatality ratio (%) | 0.05 |  |  | 8 |
| Transmission rate (β) | Dependent on R<sub>t</sub> |  |  |  |
| Rate of symptoms development (σ) |  |  |  |  |
| Scenarios | Best | Base | Worst |  |
| Effective reproductive number, R<sub>t</sub> | 1.5 | 2.5 | 3.5 | 8,14,15 |
| Test specificity (true negative rate, %) | 99.7 | 98.0 | 98.0 | 16,17 |
| Exogenous shock events (number of infections/time interval) | 0 | 5/week | 25/14 days | Assumption |
| Test characteristics | I | II | III |  |
| Sensitivity (true positive rate, %) | 70 | 80 | 90 | Assumption |
| Cost per test (\$) | 10 | 20 | 50 | Assumption |
| Time to test result return (hours) | 8 |  |  | Assumption |
| Confirmatory test sensitivity (%) | 100 |  |  | Assumption |
| Confirmatory test cost (\$) | 100 | | | Assumption |

### Compartmental Model

To the basic “susceptible-infected-removed” (or “SIR”) compartmental modeling framework, we added the following: the availability of regular, repeated screening with a test of imperfect sensitivity and specificity; creation of a new compartment for uninfected persons receiving a false positive test result; separation of the infected compartment to distinguish between undetected asymptomatics, detected asymptomatics (“true positives”), and observed symptomatics; and the importation of additional new infections via exogenous shocks (e.g., infections transmitted to students by university employees or members of the surrounding community; “super-spreader” events such as parties). We defined three epidemic severity scenarios: a “base case” with R_t_ = 2.5, a test specificity of 98%, and the exogenous introduction of five new, undetected infections into the susceptible population each week; a “worst case” with R_t_ = 3.5, a test specificity of 98%, and 25 exogenous new infections every two weeks; and a “best case” with R_t_ = 1.5, test specificity 99.7%, and no exogenous shocks.

### Isolation

We assumed that after a lag of 8 hours, individuals receiving a positive test result (true or false) and those exhibiting symptoms of COVID-19 were moved from the general population to an “isolation dormitory” where their infection was confirmed, where they were treated with supportive care, and from which no further transmissions were possible. The lag reflected both test turnaround delays and the time required to locate and isolate identified cases. Confirmed (true positive) cases remained in the isolation dormitory an average of 14 days, to ensure they were not infectious before proceeding to a recovered/immune state.^9,10^ Students with false positive results remained isolated for 24 hours, reflecting our assumption that a highly-specific confirmatory test could overturn the original diagnosis, permitting them to return to the campus population.

We assumed a symptomatic case fatality risk of 0.05% and a 30% probability that infection would eventually lead to observable COVID-19-defining symptoms in this young cohort.^8,11-13^

### Screening

We sought to evaluate both existing SARS-CoV-2 detection methods and newer technologies that could plausibly be available in the near future. Accordingly, we considered a range of different test sensitivities (70-99%), specificities (98-99.7%),^16,17^ and per test costs ($10-$50). For each combination of these test characteristics, we considered screening frequencies every 1, 2, 3, and 7 days. We assumed that a confirmatory test with 100% specificity could distinguish false positive from true positive results at a cost of $100.

### Cost-effectiveness

Next, we estimated incremental cost-effectiveness ratios, denominated in screening costs per infection averted. This measure of return on investment in screening was compared to a benchmark of value estimated by multiplying the following four terms: 1) COVID-related mortality of 0.05% in persons of college age;^8^ 2) survival loss of 60 years per college-age fatality’^20^ 3) societal willingness-to-pay (WTP) $100,000 per year of life gained;^21^ and 4) (1 + R_t_), to account for the fact that each infection averted prevents an average of R_t_ secondary infections.^8,14,15^ This method yielded a maximum WTP to avert one infection ranging from $7,500 (best case) to $10,500 (base case) to $13,500 (worst case).

Cost-effectiveness analysis identified a preferred screening strategy from among 12 possibilities – three test sensitivities (70%, 80%, and 90%) and four frequencies (1, 2, 3, and 7 times per week) – under each of the epidemic scenarios (base, worst, and best case) described above. To help decision makers understand the fiscal consequences of pursuing these preferred strategies, we also conducted a budget impact assessment, reporting the cumulative costs for the semester on a per-student basis.

## RESULTS

### Impact of Test Frequency and Sensitivity

Over an 80-day semester, in the base case, daily screening with a 70% sensitive, 98% specific test will result in 85 cumulative infections. This estimate jumps to 135/234/3,662 when tests are performed every 2/3/7 days. Raising the sensitivity of the test from 70% to 90% will reduce total infections (e.g., from 85 to 77 for daily screening and from 3,662 to 1,612 for weekly screening). But across all three epidemic severity scenarios, frequency of testing has an even more powerful impact on cumulative infections than the sensitivity of the test employed (**Figure 1**).

**Figure 1.**
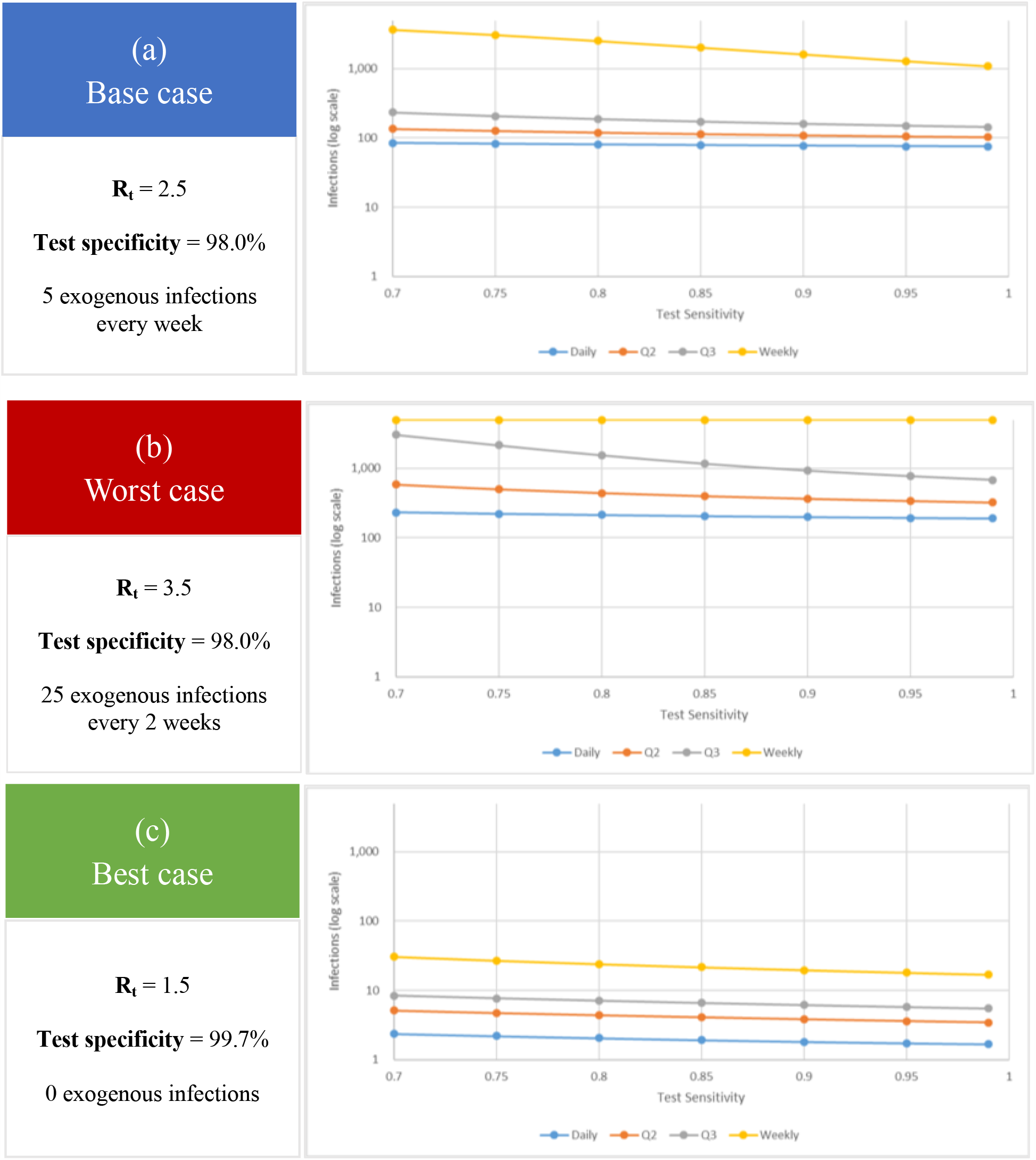
Cumulative infections as a function of test sensitivity and frequency. Over an 80-day horizon, for the (**a**) base case (R_t_ 2.5), (**b**) worst case (R_t_ 3.5), and (**c**) best case (R_t_ 1.5), these figures report cumulative infections (vertical axis; logarithmic scale) for tests with sensitivity ranging from 70-99% (horizontal axis). The colored lines denote different screening test frequencies (blue: daily screens; orange: every 2 day screen; gray: every 3 day screen; yellow: weekly screen).

### Isolation Dormitory Occupancy

In the base case (R_t_ = 2.5 and 5 exogenous infections each week), daily screening with a 70% sensitive, 98% specific test results in an average isolation dormitory census of 108 occupants, of whom 12% are truly infected (**Figure 2a**). With the frequency of screening reduced to once every 2 (3) days, overall census falls to 66 (59), as fewer tests are performed and fewer false positives are obtained; however, less frequent testing also results in greater transmission of infection and the average proportion of truly infected persons in isolation rises to 27% (46%) (**Figure 2b/2c**). Further reducing the frequency of screening to weekly causes the infected occupancy of the isolation dormitory to grow explosively (**Figure 2d**).

**Figure 2:**
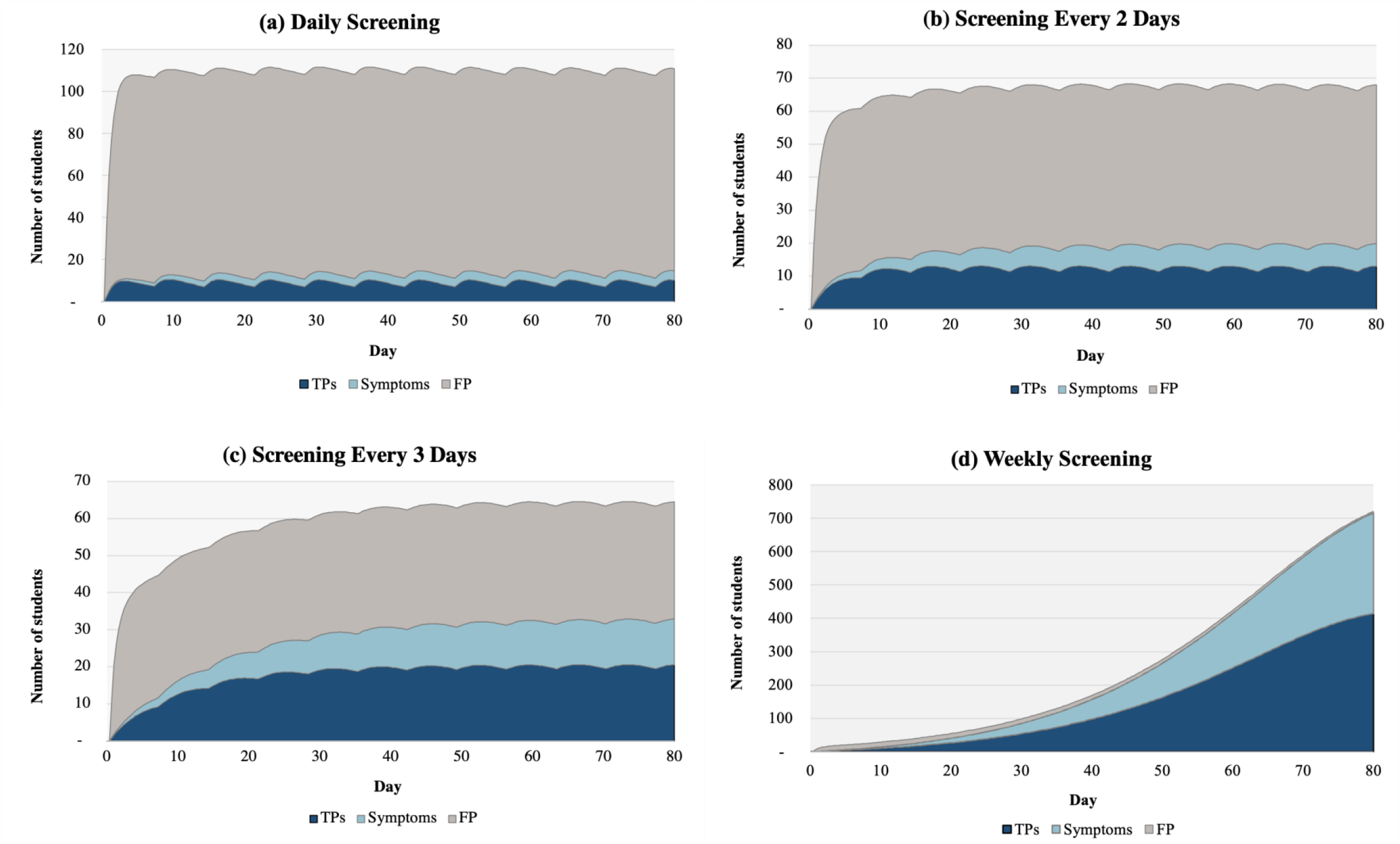
Expected daily occupancy of the isolation dormitory under base case assumptions. **Projecting the required size of the isolation dormitory**. An isolation dormitory needs to be large enough to house students with false positive results (shaded gray), students with symptoms (shaded light blue), and students without symptoms who have received true positive results (shaded dark blue). Over the 80-day horizon (time on the horizontal axis), this figure depicts the number of students in the isolation dormitory (vertical axis, note the scales are different) by indication, using a 70% sensitive, 98% specific test, under the base case scenario (Rt = 2.5). The panels show results of screening at different frequencies: (**a**) daily screening;(**b**) screening every 2 days; (**c**) screening every 3 days; and (d) weekly screening. In **Panels a through c**, the effect of exogenous shocks (5 per week) is visible in the scalloped borders; this is less evident with weekly testing where the number of true positive cases masks the comparatively small impact of exogenous shocks.

False positives – and the isolation capacity required to accommodate them – are greatly reduced using a more specific test. With daily screening in the base case, for example, increasing the test specificity from 98% to 99.7% causes the average daily census of false positives in isolation to fall from 96 to 15.

Under worst case assumptions (R_t_ = 3.5 and 25 exogenous infections every two weeks) average census grows from 127 (26% truly infected) with daily screening to 308 (92% truly infected) with screening every 3 days (**Figure S2**). With weekly screening, virtually the entire student population will have been infected before the 80-day semester is concluded.

In the best case (R_t_ = 1.5, no exogenous shocks, and a 99.7% specific test), average occupancy of the isolation dormitory is light (5 infected, 2 false positives) and can be controlled with no more than weekly screening (**Figure S3**).

### Cost-effectiveness and budget impact

In the base case, screening with a less expensive, less sensitive test dominates (i.e., costs less and averts greater numbers of infection) screening with more expensive, more accurate tests for all plausible WTP values. At the benchmark maximum WTP ($10,500/infection averted in the base case), screening every 2 days with a 70% sensitive test is the preferred strategy. If WTP exceeds $46,400 per infection averted, daily screening with this same test is preferred (**Table 2**). Under worst-case assumptions, daily screening strategies are the only undominated choices for all WTP values exceeding $6,600/infection averted; at the benchmark maximum WTP ($13,500/infection averted in the worst case), daily screening with the least sensitive (70%) test is the preferred choice. Under best-case assumptions (WTP maximum $7,500 per infection averted), weekly screening with a 70% sensitive test is preferred.

**Table 2:** Results of the incremental cost-effectiveness analysis ($/COVID-19 infection averted) in the base (top), worst (middle) and best (bottom) case scenarios. Preferred strategies at the maximum willingness-to-pay (WTP) threshold are shaded gray.

| Frequency | Test Sensitivity (%) | Cost (\$) | Total Infections | Incremental Cost-effectiveness Ratio (\$/infection averted)* |
| --- | --- | --- | --- | --- |
| **Base Case Scenario ($R_t$ 2.5, 5 exogenous shock infections each week)** |  |  |  |  |
| **Maximum willingness-to-pay = \$10,500/infection averted** | | | | |
| Weekly | 70 | 718,700 | 3,662 | ( - ) |
| Weekly | 80 | 1,210,700 | 2,525 | dominated |
| Every 3 days | 70 | 1,567,600 | 234 | 200 |
| Every 2 days | 70 | 2,350,300 | 135 | 7,900 |
| Weekly | 90 | 2,776,700 | 1,612 | dominated |
| Every 3 days | 80 | 2,870,000 | 187 | dominated |
| Every 2 days | 80 | 4,306,700 | 120 | dominated |
| Daily | 70 | 4,664,300 | 85 | 46,400 |
| Every 3 days | 90 | 6,781,400 | 160 | dominated |
| Daily | 80 | 8,550,800 | 81 | 852,300 |
| Every 2 days | 90 | 10,177,700 | 109 | dominated |
| Daily | 90 | 20,211,100 | 77 | 3,480,900 |
| **Worst Case Scenario ($R_t$ 3.5, 25 exogenous shock infections every 2 weeks)** |  |  |  |  |
| **Maximum willingness-to-pay = \$13,500/infection averted** | | | | |
| Weekly | 70 | 548,200 | 4,991 | ( - ) |
| Weekly | 80 | 842,100 | 4,992 | dominated |
| Every 3 days | 70 | 1,490,400 | 3,052 | dominated |
| Weekly | 90 | 1,714,600 | 4,990 | dominated |
| Every 2 days | 70 | 2,297,000 | 584 | 400 |
| Every 3 days | 80 | 2,701,400 | 1,545 | dominated |
| Every 2 days | 80 | 4,212,900 | 442 | dominated |
| Daily | 70 | 4,613,200 | 233 | 6,600 |
| Every 3 days | 90 | 6,443,300 | 929 | dominated |
| Daily | 80 | 8,456,600 | 213 | 194,800 |
| Every 2 days | 90 | 9,969,000 | 366 | dominated |
| Daily | 90 | 19,990,000 | 199 | 845,900 |

\*Dominated strategies are those that cost more and result in more infections than some combination of other strategies.

| Frequency | Test Sensitivity (%) | Cost (\$) | Total Infections | Incremental Cost effectiveness Ratio (\$/infection averted)* |
| --- | --- | --- | --- | --- |
| **Best Case Scenario ($R_t$ 1.5, no exogenous shocks, 99.7% specific test)**<br>**Maximum willingness-to-pay <\$7,500/infection averted** | | | | |
| **Do Nothing** | - | 0 | 1,371 | ( - ) |
| **Weekly** | 70 | 586,600 | 31 | 400 |
| **Weekly** | 80 | 1,154,800 | 24 | dominated |
| **Every 3 days** | 70 | 1,368,400 | 8 | 35,200 |
| **Every 2 days** | 70 | 2,051,900 | 5 | 209,200 |
| **Every 3 days** | 80 | 2,696,400 | 7 | dominated |
| **Weekly** | 90 | 2,860,300 | 20 | dominated |
| **Every 2 days** | 80 | 4,043,700 | 4 | dominated |
| **Daily** | 70 | 4,098,500 | 2 | 742,600 |
| **Every 3 days** | 90 | 6,680,500 | 6 | dominated |
| **Daily** | 80 | 8,077,500 | 2 | dominated |
| **Every 2 days** | 90 | 10,019,100 | 4 | dominated |
| **Daily** | 90 | 20,014,700 | 2 | dominated |

\*Dominated strategies are those that cost more and result in more infections than the next least costly strategy.

Over the 80-day semester, the per-student costs of implementing the preferred screening strategy will be $120, $470, and $920 in the best, base, and worst case scenarios, respectively (**Table 3**).

**Table 3:** Per student costs for optimal policies over an 80-day horizon under base, worst, and best case scenarios

| Scenario | Optimal Policy | Cost per Student (\$) |
| --- | --- | --- |
| **Base case ($R_t = 2.5$)** | Screening every 2 days, 70% sensitivity | 470 |
| **Worst case ($R_t = 3.5$)** | Daily screening, 70% sensitivity | 920 |
| **Best case ($R_t = 1.5$)** | Weekly screening, 70% sensitivity | 120 |

## DISCUSSION

The safe return of students to residential colleges demands an effective SARS-CoV-2 monitoring strategy. We find that a highly specific screening test that can easily be administered to each student every one to seven days – and that reports results quickly enough to permit newly detected cases to be isolated within hours – will be sufficient to blunt the further transmission of infection and control outbreaks at a justifiable cost.

Of the many uncertain variables driving our assessment of the required frequency of screening, we highlight the effective reproductive number, R_t_. This uncertain measure of the transmission potential of infection will depend in part on factors that are within the control of students and university administrators. Strict adherence to hand-washing, mask-wearing, public space occupancy limits, and other best practices could drive R_t_ down to best-case levels, rendering containment controllable with testing as infrequent as weekly. However, any relaxation of these measures in the residential college setting could easily drive R_t_ to worst-case levels, requiring screening as frequent as daily. All members of the university community must understand the fragility of the situation and the ease with which inattention to behavior may propagate infections and precipitate the need once again to shut down campus.

Much depends on the judicious management of positive test results, both true and false. Rapid detection, confirmation, isolation, and treatment of true positives is, of course, essential. We find that frequent screening with a test of modest sensitivity and a turnaround time to results of 8 hours will be sufficient for this purpose. The greater difficulty lies in managing the overwhelming number of false positives that will inevitably result from repeated screening for low-prevalence conditions. False positive results threaten to overwhelm isolation housing capacity, a danger whose gravity increases with screening frequency. The specificity of the initial screen will matter far more than its sensitivity.

Even with a 98% specific screening test, false positives will present a challenge. Until a confirmatory test result is obtained, anyone receiving a positive test result will be presumed to be infectious and needing to be separated from other students. Setting aside the logistical challenges and financial costs, administrators must anticipate the anxiety such separations may provoke among both students and their families. Excessive numbers of false positives may fuel panic and undermine confidence in the reliability of the monitoring program. It may be possible to work with test manufacturers to tune test kits for use in this setting, sacrificing some small measure of sensitivity in favor of higher specificity.

Obtaining an adequate supply of testing equipment will be a challenge. On a college campus of 5,000 enrollees, screening of the students alone every two days will require roughly 195,000 test kits over the abbreviated semester. Our analysis assumed per test costs (including the test equipment and associated personnel costs) ranging from $10-$50. Lower-cost, self-administered testing modalities may soon be available and could make screening more affordable. Pooling could also facilitate more efficient, higher volume screening.^22^ However, pooling introduces its own logistical challenges and could increase the time to definitively identify and isolate a positive case, resulting in further transmission and provoking anxiety among the many uninfected students notified that they are among the members of an initially positive pool.

We have tried to help decision makers make sense of the “value” question by conducting a cost-effectiveness analysis and by comparing our findings to a rough estimate of the societal willingness to pay per infection averted.^23^ While we have adhered to the broad outlines of recommended practice for the conduct of economic evaluation,^23^ we urge readers to interpret our results with caution. The majority of our assumptions are conservative – that is, they understate the value of more frequent testing. For example, we ignore the clinical harms and attributable costs of COVID-19-related morbidity and treatment. We also ignore the value of infections averted beyond the student population. However, a few assumptions (e.g., our failure to account for the economic and quality of life effects of false positives) may pull in the direction of less testing.

The simple model underlying this analysis has notable limitations. We assumed homogenous mixing without age-dependent transmission. We did not explicitly include the impact of screening on faculty and staff, though we did allow for the importation of infections from exterior sources. We assumed that no students arrive on campus with immunity to COVID-19. Finally, we excluded the impact of symptom screening and contact tracing. Given that both are logistically challenging, this is a noteworthy omission; our results suggest that with frequent enough screening, neither symptom checking nor contact tracing would be necessary for epidemic control.

Reopening college imposes risks that extend beyond students to the faculty who teach them, to the many university employees (administrative staff, dining hall workers, custodians) who come into close daily contact with them, and to the countless other members of the surrounding community with whom they come into contact. University presidents have a duty to consider the downstream impact of their reopening decisions on these constituencies. However, their first responsibility is to the safety of the students in their care. So, while we certainly do not intend to minimize the broader effects of the reopening decision, we have quite deliberately excluded from consideration any transmissions exported off campus.

We believe there is a safe way for students to return to college in the Fall of 2020; the question is whether it is feasible today on a large scale. Coupled with strict behavioral interventions that keep R_t_ below 2.5, a rapid, inexpensive and even poorly sensitive (>70%) test, conducted at least every 2 days, would produce a modest number of containable infections and would be cost-effective.

## Data Availability

All data used in this analysis were obtained from published, publicly available sources, which we have cited in the manuscript.

## Supplementary Appendix

**Testing for COVID-19 in higher education:**

**What testing do we need to open college campuses?**

## Model description

We developed a dynamic, compartmental model using a modified “susceptible-infected-recovered” (or SIR) framework. The model portrays the epidemiology and natural history of infection in a homogeneous population of at-risk individuals as a sequence of transitions, governed by difference equations, between different health states (or “compartments”). The flow diagram (**Figure S1**, below) illustrates the modifications we made to the basic SIR framework:

- Addition of regular, repeated screening with a test of imperfect sensitivity and specificity.
- Removal of infected individuals from the transmitting population based on either screening test findings or the development of COVID-defining symptoms.
- Removal (and return) of uninfected individuals from the transmitting population based on “false positive” screening test findings.
- Importation of additional new infections from exogenous sources (e.g., infections transmitted to students by university employees or members of the surrounding community.

### Compartments

We defined a total of 7 model compartments, divided into three pools:

- Active transmission and testing pool. Everyone is in this pool at time 0. All transmission of infection takes place between individuals in this pool. This is also the pool in which screening for infection takes place.
  - U: Uninfected, susceptible individuals
  - A: Infected, asymptomatic
  Note that, without testing, individuals in these two compartments are indistinguishable from one another.
- Isolation pool. Individuals in this pool are assumed to be isolated from the active transmission pool and from one another. It is assumed that transmission is not possible within this pool.
  - S: Infected, symptomatic (true) positive test result
  - TP: Infected, asymptomatic, (true) positive test result
  - FP: Uninfected, false positive result
- Removed pool. Individuals in this pool are assumed to play no role either in the transmission of infection or in testing activities.
  - R: Recovered
  - D: Dead

## Parameters

β: rate at which infected individuals contact susceptibles and infect them

τ: rate at which individuals in the testing pool are screened for infection

δ: rate at which individuals in the symptomatic compartment die

ρ: rate at which infected individuals recover from disease and are removed

σ: rate of symptom onset for infected individuals

μ: rate at which false positives are returned to the Uninfected compartment

Se: sensitivity of the screening test

Sp: specificity of the screening test

I(t): an indicator function which assumes value 1 if an exogenous shock takes place in cycle t; 0 otherwise

X: number of imported infections in a given exogenous shock

The model uses a cycle time of 8 hours. All rates are calculated per 8-hour cycle.

### Governing equations

- Uninfected (t+1) = Uninfected (t) – New Infections – New FPs + Returning FPs – Exogenous Shocks

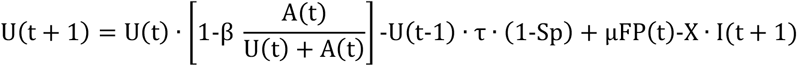
- Asymptomatic (t+1) = Asymptomatic (t) –symptoms - recoveries+ New Infections – TPs + Exogenous Shocks

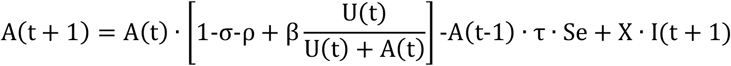
- False Positives (t+1) = False Positives (t) – Returning FP+ New FPs

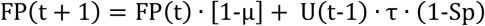
- True Positives (t+1) = True Positives (t) – Symptoms – Recovery + New TPs

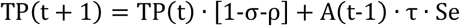
- Symptomatic (t+1) = Symptomatic (t) – Recovery – Mortality + New Symptoms

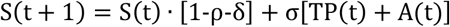
- Recovered (t+1) = Recovered (t) + New Recoveries

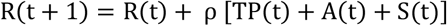
- Deaths (t+1) = Deaths (t) + New Deaths

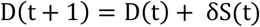
- N = U + A + S + TP + FP + R + D = Total population size (constant)

Note that there is a lag of one cycle between the time that a test is conducted and the time that persons receiving a positive test result are moved to the isolation pool.

### Initial conditions

U(0) = 4,990

A(0) = 10

All other compartments are empty at time 0.

### Estimating Key Rate Parameters

1. <u>σ: rate of symptom onset for infected individuals</u>. We assumed that 30% of all infected individuals would eventually develop symptoms. In the absence of a screening program, this implies that σ / (σ+ρ) = 0.3. Assuming a mean recovery time of 14 days and computing all rates per 8-hour cycle yields ρ = 1/(3* 14 days) and we solve for σ = 0.0102.
2. <u>β: rate at which infected individuals contact susceptibles and infect them.</u>The effective reproductive number R_t_ = β / (σ+ρ). We assumed Rt = {1.5, 2.5, 3.5}, which implies β = {0.051, 0.085, 0.119}. Recall that all rates are estimated per 8-hour cycle.
3. <u>δ: rate at which individuals in the symptomatic compartment die.</u> We assumed that the symptomatic case fatality risk was 0.05%. This implies [σ / (σ+ρ)]* [δ / (δ+ρ)] = 0.0005 and permits us to solve for δ = 0.00004.

**Figure S1.**
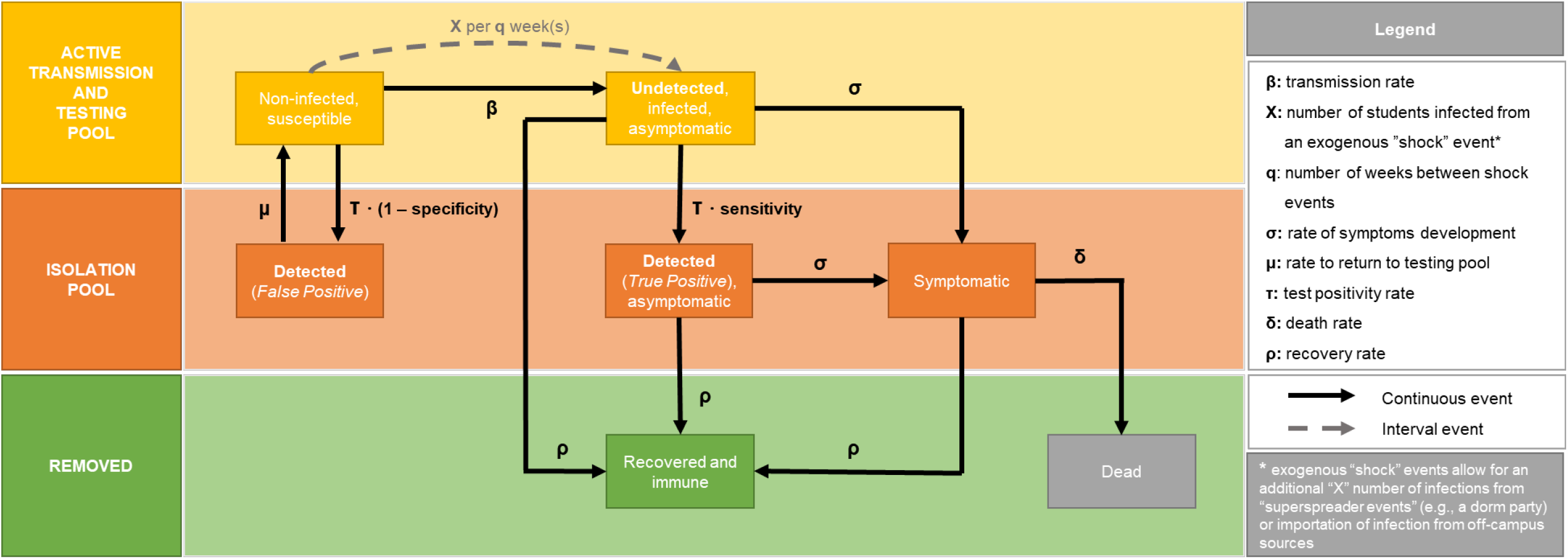
Model schematic and input parameters

**Figure S2:**
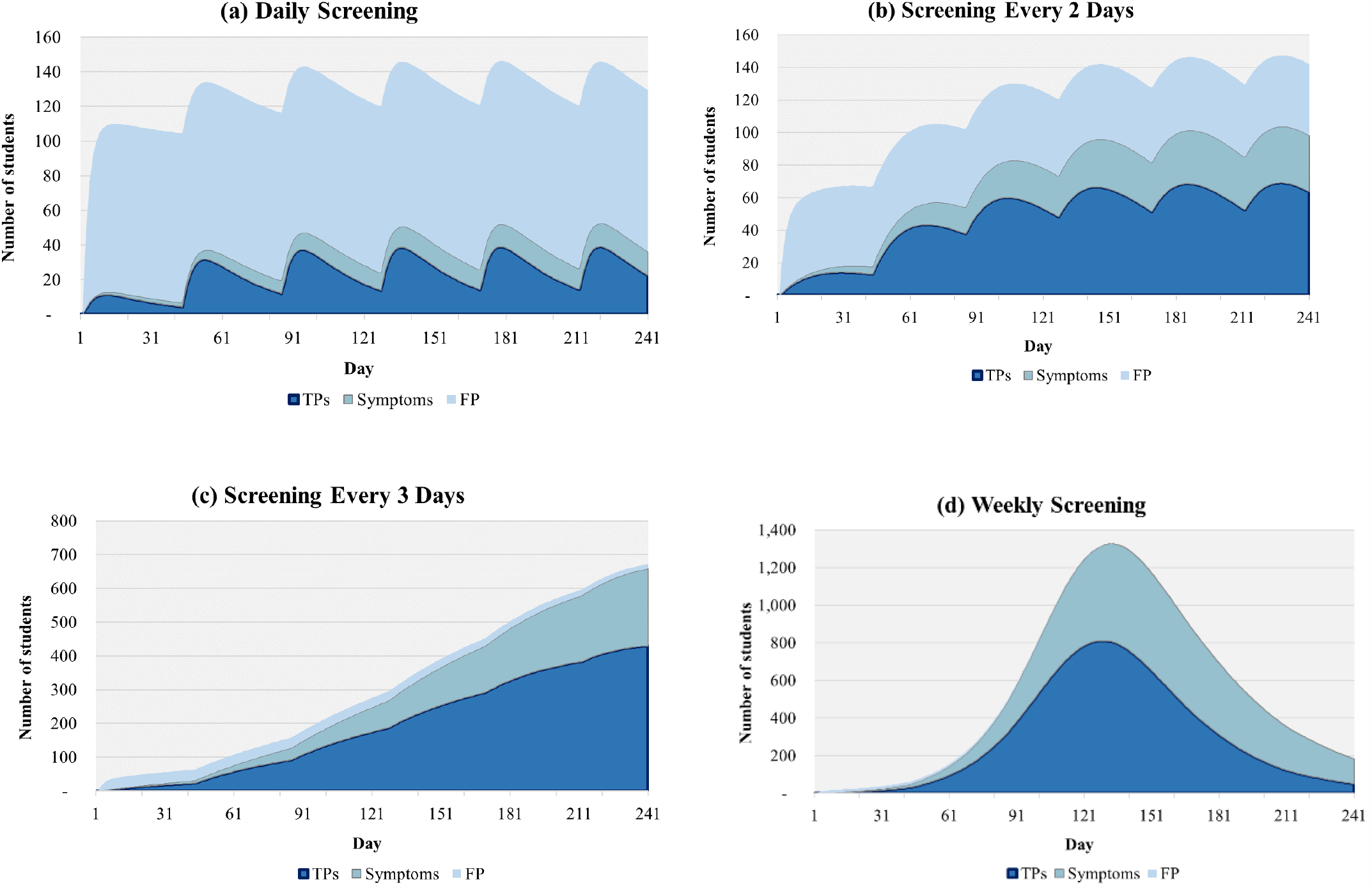
Expected daily occupancy of the isolation dormitory under worst case assumptions.

**Figure S3:**
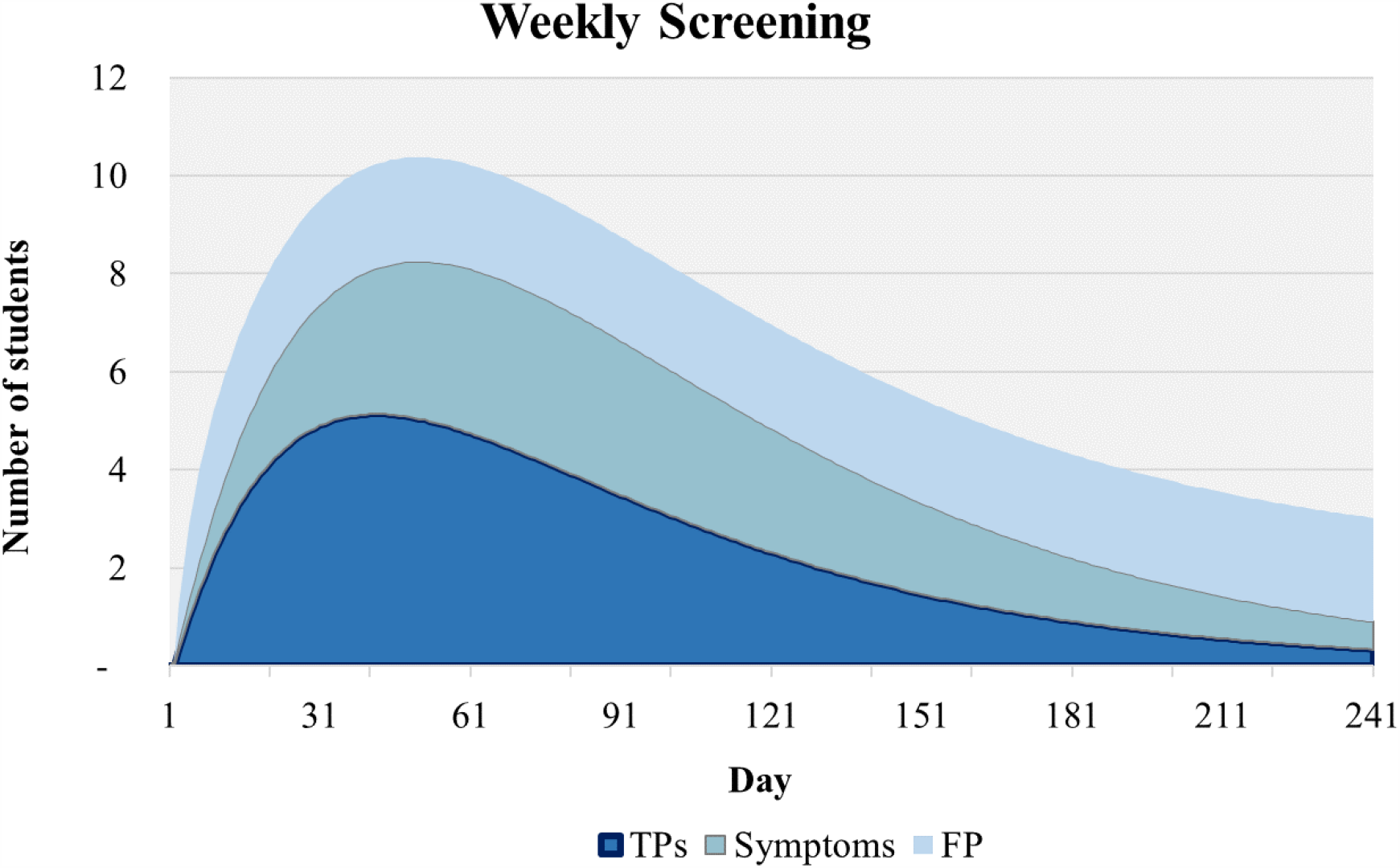
Expected daily occupancy of the isolation dormitory under best case assumptions.

